# Disproportionate COVID-19 Related Mortality Amongst African Americans in Four Southern States in the United States

**DOI:** 10.1101/2020.06.08.20124297

**Authors:** Nicole Betson, Anirban Maitra

**Affiliations:** Department of Translational Molecular Pathology, The University of Texas MD Anderson Cancer Center, Houston, Texas

**Keywords:** COVID-19, African American, Mortality

## Abstract

**Background:** African Americans have been severely affected by COVID-19 noted with the rising mortality rates within the African American community. Health disparities, health inequities and issues with systemic health access are some of the pre-existing issues African Americans were subjected to within the southern states in the United States. Second, social distancing is a critical non-pharmacological intervention to reduce the spread of COVID-19. However, social distancing was not practical and presented a challenge within the African American community, specifically, in the southern states.

**Objective:** This article assesses the effect of COVID-19 on African Americans in the southern states.

**Methodology:** This short communication queried the publicly available Department of Health statistics on COVID-19 related mortality and underlying health conditions in four southern states (Alabama [AL], Georgia [GA], Louisiana [LA] and Mississippi [MS]) with a high proportion of African American residents. Second, the unacast COVID-19 toolkit was used to derive a social distancing (SD) grade for any given state, based on three different metrics: (i) percent change in average distance travelled (ii) percent change in non-essential visits and (iii) decrease in human encounters (compared to national baseline).

**Results:** Across the four states, on average, as many as 54% of COVID-19 related deaths are in the African American community, although this minority group comprises only 32% of the population cumulatively. This article finds that all four southern states received a social distancing grade of F. COVID-19 have demonstrated that adverse outcomes are higher in individuals with underlying health conditions such as diabetes, cardiovascular diseases, or pre-existing pulmonary compromise.

**Conclusion:** The COVID-19 pandemic has exposed racial disparities in our healthcare system that disproportionately impacts African Americans within the four states of the southern United States. In addition, the lack of diversity in the healthcare system likely impacts this disproportionate impact on African American communities because it is not able to address its primary obligations within minority communities. Recognizing that there is a great need for African American representation or diversity in the health workforce would be able to better address the health disparities.

---

We have read with great interest the recent series of commentaries in CDC on disparities in hospitalization and death rates amongst African American populations compared to Caucasians ^1-3^. However, a systematic quantitative assessment of these important metrics, critical for implementing public health measures like testing, tracing and isolation in the highest risk communities, is still lacking.

We queried the publicly available Department of Health (DOH) statistics on COVID-19 related mortality in four southern states (Alabama [AL], Georgia [GA], Louisiana [LA] and Mississippi [MS]) with a high proportion of African American residents. Across the four states, on average, as many as 54% of COVID-19 related deaths are in the African American community, although this minority group comprises only 32% of the population cumulatively. Specifically, the proportion of African Americans residing in a given southern state and the proportion of COVID-19 related mortality that are comprised of African Americans in that state are as follows: AL – 26.6% and ‘50%, GA – 31.5% and ∼50%, LA – 32.2% and 59%, and MS – 37.7% and 57%. The number of deaths in the minority population has kept at par with Caucasians in three of the four states, and exceeded the latter in the hardest hit state of LA (**Figure 1**).

**Figure 1:**
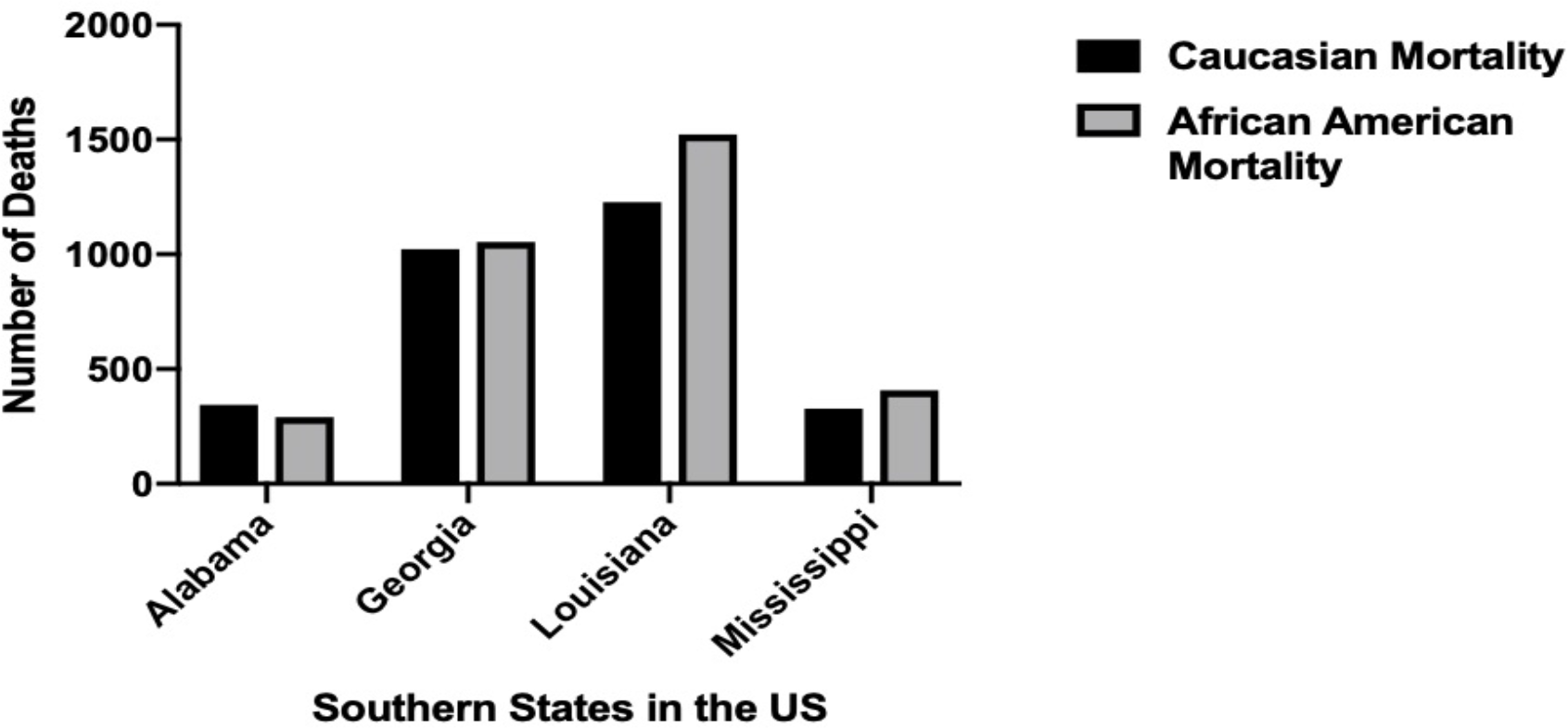
comparative Numbers of COVID-19 Related Mortality for caucasions and Africans Americans in Four Southern States of the United States.

There are likely to be several reasons for the disproportionate causes of mortality amongst African Americans in the four southern states we have interrogated here. First, emerging clinical series on COVID-19 have demonstrated that adverse outcomes are higher in individuals with underlying health conditions such as diabetes, cardiovascular diseases, or pre-existing pulmonary compromise ^4,5^. Indeed, in three of the states (AL, LA and MS) where such data is publicly cataloged, many of the patients dying from causes related to confirmed COVID-19 had an underlying chronic health condition **(Table 1)**. Secondly, social distancing is a critical non-pharmacological intervention to reduce the spread of COVID-19. The unacast COVID-19 toolkit (https://www.unacast.com/covid19/social-distancing-scoreboard) can be used to derive a social distancing (SD) grade for any given state, based on three different metrics: (i) percent change in average distance traveled (ii) percent change in non-essential visits and (iii) decrease in human encounters (compared to national baseline), each of is an average to create the overall grade for that state. Based on these parameters, all four states received a SD grade of F. Unfortunately, the suboptimal SD measures adopted statewide is even more likely to impact individuals living in crowded urban localities, those relying on public transport, or families where one or more members have been deemed as “essential” at-work personnel. According to PovertyUSA (www.povertyUSA.org), the poverty rates in the four states stands at 18.4% on average, which is greater than the national rate of 14.1%; in addition, of the reported overall numbers, the average poverty rate amongst the African American community (29.1%) is approximately 2.3 times higher compared to Caucasians (12.6%). The median household income reported by US Census Bureau for AL, GA, LA and MS were $49,861, $58,756, $47,905, and $44,717, respectively, which is, on average, more than $11,600 less than the national median household income ($61,937).

**Table 1.** Underlying Medical Conditions in Confirmed COVID-19 Cases or Deaths

| State | Underlying Medical Conditions | Laboratory Confirmed COVID-19 Death |  |
| --- | --- | --- | --- |
|  |  | Number of Cases | Percentage of Cases |
| AL | Chronic Lung Disease | 98 | 24.9% |
|  | Diabetes Melitus | 154 | 39.2% |
|  | Cardiovascular Disease | 249 | 63.4% |
| LA | Chronic Lung Disease | N/A | 11.2% |
|  | Diabetes Melitus | * | 34.7% |
|  | Cardiovascular Disease |  | 32.4% |
| GA** | Chronic Lung Disease | 3446 | 6.87% (Confirmed Cases) |
|  | Diabetes Melitus | 5055 | 10.1%(Confirmed Cases) |
|  | Cardiovascular Disease | 5854 | 11.7% (Confirmed Cases) |
| MS | Chronic Lung Disease | 109 | 13.3% |
|  | Diabetes Melitus | 215 | 26.3% |
|  | Cardiovascular Disease | 225 | 27.5% |

\* In LA, the specific numbers for comorbidities associated with COVID-19 related deaths are not reported, only the percentages.
\*\* In GA, comorbidity status is only available for confirmed COVID-19 cases, and not for COVID-19 related deaths
AL, https://www.alabamapublichealth.gov/covid19/assets/cov-al-cases-042820.pdf
LA, http://ldh.la.gov/coronavirus/
GA, https://dph.georgia.gov/covid-19-daily-status-report
MS, https://msdh.ms.gov/msdhsite/_static/14,0,420.html
Data accessed on June 7<sup>th</sup>, 2020.

While we cannot exclude an underlying genetic basis to the disproportionate burden of mortality related to COVID-19 among the African American community (for example *ACE2* polymorphisms), the emerging data strongly suggests that African American communities face an array of social and economic inequities that impact their baseline health status and access to healthcare, and is likely driving the course of the pandemic in this population ^6^. Measures at mitigating the impact of COVID-19 on African Africans need to address these inequities in lockstep with more conventional interventions.

## Author Statement

Ethical approval was not necessary because this study did not involve human subjects and animal studies

## Data Availability

This short communication queried the publicly available Department of Health statistics on COVID-19 related mortality and underlying health conditions in four southern states.

https://jamanetwork.com/journals/jama/fullarticle/2764789

https://jamanetwork.com/journals/jama/fullarticle/2766098

https://jamanetwork.com/journals/jama/fullarticle/2766096

https://www.cdc.gov/mmwr/volumes/69/wr/mm6918e1.htm

https://www.nejm.org/doi/full/10.1056/NEJMc2010419

https://www.ncbi.nlm.nih.gov/pmc/articles/PMC7166096/

